# Reduction in maternal mortality ratio varies by district in Sidama Regional State, southern Ethiopia: Estimates by cross-sectional studies using the sisterhood method and a household survey of pregnancy and birth outcomes

**DOI:** 10.1101/2022.10.02.22280613

**Authors:** Aschenaki Zerihun Kea, Bernt Lindtjorn, Achamyelesh Gebretsadik, Sven Gudmund Hinderaker

## Abstract

**Introduction:** Maternal mortality studies conducted at national level do not provide subnational and local estimates useful for program planning and monitoring maternal mortality at lower administrative level. The aim of this study was to estimate life time risk (LTR) of maternal death and maternal mortality ratio (MMR), assess variations and reduction in MMR in Sidama Regional State, southern Ethiopia.

**Methods:** This is cross-sectional study employed the sisterhood method as part of larger household maternal mortality survey that used a 5-year recall of pregnancy and birth outcomes. We visited 8880 households and interviewed 17444 respondents 15-49 years age. We assessed reduction in MMR using the sisterhood method (main focus of this paper) and a household survey of pregnancy and birth outcomes.

**Result:** We analysed 17374 (99.6%) respondents; 8884 (51.1%) men and 8490 (48.9%) women. The 17,374 respondents reported 64,387 maternal sisters who had reached reproductive age. Among these, 2,402 (3.7%) sisters had died; 776 (32.3%) were pregnancy related deaths. The LTR of maternal death was 3.2% and MMR was 623 (95% CI: 573-658) per 100000 live births (LB), with reference to year 2010. The remote district (Aroresa) had a MMR of 1210 (95% CI: 1027-1318) per 100,000 LB. Assessment of MMR reduction using the two maternal mortality estimation methods showed that the MMR in Sidama Regional State declined over the past years. Significant reduction in MMR was observed in districts located near the regional centre. However, no reduction was observed in districts located distant to the regional centre.

**Conclusion:** The high MMR with district level variations and lack of mortality reduction in districts located distant to the centre highlights the need for instituting interventions tailored to the local context to save mothers and accelerate reduction in MMR.

## Introduction

The government of Ethiopia has been implementing interventions aiming to reduce maternal mortality since the commencement of the Millennium Development Goal (MDG) [1], and the maternal deaths in the country fell by 61% during the MDG period. Despite these achievements, in 2017, there were an estimated 14000 maternal deaths with corresponding maternal mortality ratio (MMR) 401 per 100,000 live births (LB).

Currently, maternal deaths are not routinely tracked using vital registration in Ethiopia [2, 3]. In certain areas, maternal deaths are tracked through studies conducted at special settings like demographic surveillance sites [4] and hospitals [5]. The data used for monitoring maternal mortality in the country largely comes from Demographic and Health Surveys (DHS) [6] and UN agency studies [7] conducted periodically at national level. However, studies conducted at national level do not provide data on magnitude and variations of maternal deaths at district level. Hence, at lower administrative level in Ethiopia, it is difficult to understand the magnitude and variations of maternal deaths and ensure reduction of MMR.

A recent population based household survey conducted in Sidama Regional State identified a high MMR with district level variations. The overall MMR of the region, reported by this study was 419 (95% CI: 260-577) per 100,000 LB, but Aroresa district (the remote district) had a MMR 1142 (95% CI: 693-1591) deaths per 100,000 LB [8].

We carried out a population based household survey employing the indirect sisterhood method to measure maternal mortality in Sidama Regional State, southern Ethiopia. The indirect sisterhood method was part of larger maternal mortality survey aimed at measuring maternal mortality in Sidama Regional State using a retrospective 5-year recall period of pregnancy [8]. The larger study also included measuring physical access to skilled delivery using Geographic Information System. The 5-year pregnancy and birth outcome household survey helps to understand the magnitude of and district level variations in maternal mortality for recent years, whereas the indirect sisterhood method of this paper is used to understand past maternal mortality level and variations in the region and in districts included in the study.

The indirect sisterhood method has been validated and used in different settings [9-12]. Our specific objectives in this study using the indirect sisterhood method were 1) to estimate life time risk (LTR) of maternal death and corresponding MMR; 2) to assess variations in maternal mortality estimates based on characterstics of the respondents; 3) to assess reduction in MMR in Sidama Regional State over the past year by using MMR of the sisterhood method (main focus of this paper) with results from the 5-year recall of pregnancy and birth outcome household survey.

## Methods

### Study design and setting

We carried out a cross sectional study employing the sisterhood method (main focus of this paper) as part of larger maternal mortality household survey that used a retrospective 5-year recall of pregnancy and birth outcomes [8]. The study was conducted in Sidama Regional State, Southern Ethiopia. Six districts (Aleta Chuko, Aleta Wondo, Aroresa, Daela, Hawassa Zuriya and Wondogenet) of Sidama Regional State were included in the study. The study was conducted from July 2019 to May 2020. Sidama Regional State is one of 11 regional states in the country, and its capital Hawassa is located 273 km south of Addis Ababa. The population of the region projected for 2020 was 4.3 million people. Administratively, the region is divided into 30 rural districts (*woredas*), 6 town administrations and 536 rural *kebeles* (the smallest administrative structure with an average population of 5000 people). Under the *kebele*, there are local structure known as *limatbudin* (local administrative unit consisting of 40-50 neighbouring households in average).

### The sisterhood method

The sisterhood method utilizes information obtained from adult men and women about the deaths of their sisters born to the same mother, and who were in reproductive age. The sisterhood method uses the proportions of adult sisters dying during pregnancy, childbirth, or the puerperium reported by adults during a census or survey. In this way, a cohort of women in reproductive age at risk of pregnancy related death is created [12, 13]. The LTR of maternal death is estimated from sister unit of risk exposure to maternal mortality. MMR is approximated using total fertility rate (TFR) of the study area [12, 13]. The MMR estimate from the indirect sisterhood method in average refers to 10-12 years prior to data collection and even can extends to 35 years when the respondents are older [12].The sisterhood method is acclaimed for simplicity, minimal time and inexpensiveness. However, it’s not appropriates for monitoring short term progress [14].

### Study population and sampling techniques

Men and women aged 15-49 years in Sidama Regional State were source population of the study, and men and women of same age and residing in sampled households were the study population.

The detailed sampling techniques of the study has been described elsewhere [8]. Below, we summarize the sampling technique of the sisterhood method conducted as part of a larger maternal mortality household survey. Multistage cluster sampling techniques was employed to identify the study participants. At first stage, we randomly selected six districts from the region. At second stage, we selected 40 *kebeles* from the six districts proportional to the size of *kebeles* in the districts. Thirdly, from each *kebele*, we sampled 6 *limatbudin* and finally selected 37 households from each *limatbudin*. Men and women 15-49 years age residing in the sampled households were eligible for the sisterhood interview. When two or more men or women born to the same mother were found in the same household, one of them was selected by a lottery method to avoid multiple counting of the same sisters.

### Variables

Life time risk of maternal deaths and MMR; maternal deaths per 100,000 LB were the outcome measures of the study. We also collected characterstics of the respondents, like sex, age and educational level to assess their association with the outcome measures of interest.

### Data source and measurement

We carried out an interview with 17444 participants and analysed the data of 17374 participants; 8884 (51.1%) men and 8490 (48.9%) women 15-49 years age. The participants were residents of the 8880 households of the survey [8]. The data was collected by diploma level teachers recruited from each *kebele* who were familiar with the culture and language of the study area. The data collectors conducted an interview with men and women 15-49 years using interviewer administered questionnaire. Participants were asked the following four standard questions used in indirect sisterhood method [12]. 1) How many sisters (born to the same mother) have you ever had who reached 15 years? 2) How many of these sisters who ever-reached 15 years are alive now? 3) How many of these sisters who ever-reached 15 years are dead? 4) How many of these dead sisters died while they were pregnant, or during childbirth, or during the six weeks after the end of pregnancy?

The first question addresses the number of sisters who were at risk of pregnancy related deaths while the fourth question identifies the actual number of pregnancy related deaths. Interviewers were observant to ensure the sum of question 2 and 3 equals to question 1 as a quality assurance of numbers. The period of six weeks after the end of pregnancy in question 4 was approximated 2 months after the end of pregnancy.

### Data quality control

We adapted the standard sisterhood questions to local context [12]. The questionnaire was prepared in English, translated into local language (*Sidaamu Afoo*) and back translated to English by another individual to check its consistency. Before actual data collection, the questionnaire was pretested in one district not included in the study.

The data collectors and supervisors were trained by the principal investigator. The training was part of the larger survey training in which one day was scheduled for the sisterhood. The training content included discussion on the content and aim of each question, interview techniques and role plays. The data collectors were supervised by two public health officers recruited from each district who were also familiar with the culture and language of the study setting. The supervisors followed the data collectors, checked consistency and completeness of the questionnaire. When eligible respondents were absent during initial visit, the data collectors revisited the households the next day.

The data was double entered using EpiData software (EpiData Association 2000-2021, Denmark). Consistency of the two entries was checked and discrepancies validated from the hard copies of the questionnaire.

### Sample size determination

Hanley and colleagues suggested that, in settings where the MMR within the range of 500 per 100,000 LB, the death of 385 sisters with (± 10% margin of error) will be reported from interviews of 13000 adult respondents aged 15-49 [13]. According to the 2016 EDHS report, the MMR of Ethiopia was 412/100,000 LB [6]. Adding 10% non-responses, we decided to interview 14,300 respondents. The number of households planned for the larger maternal mortality household survey was 8880. We assumed that, surveying the 8880 household would be sufficient to get the desired sample of adult respondents for the sisterhood study.

### Statistical analysis

Stata 15 were used for data analysis. Before LTR and MMR computation, two adjustments were made [12]. The first adjustment was made to the younger age group. The respondents in younger age group (15-19 and 2024) would have sisters who have not yet reached age 15. Hence, we created hypothetical number of sisters that would have been reported by the young age group. To get this number, we calculated the average number of sisters reported by older age group (25-49) and multiplied by the initial number of sisters reported by the younger age group. The average number of sisters reported by age group 25-49 was 2.77 (37072/13401). Therefore, the number of sisters reported by age group 15-19 was 1475 and multiplied by 2.77 it equals to 4086; and same for age group 20-24 (8386 sisters x 2.77=23229).

The second adjustment was made to get sisters exposed to full lifetime risk exposure [12]. As we interviewed participants aged below 50, we did not have information about full lifetime risk of all their sisters. Hence, we multiplied the sister unit of risk exposure of each age group with the adjustment factor in order to get “complete” sister units of risk exposure. The adjustment factor was a number calculated for risk exposure of different age group in developing countries which is independent of a particular country [12].

Descriptive statistics with means and percentages were computed to describe participants’ characteristics. The LTR for maternal death was obtained by dividing the total number of maternal deaths reported by the estimated total number of sisters exposed (LTR= (Total number of maternal deaths)/ (Total sister units of risk exposure). We used total fertility rate (TFR) of rural population of Ethiopia 5.2 [6] to estimate MMR from the LTR. (MMR = (1-[(1-LTR) 1/TFR] × 100,000), using formulas specified by Hanley et al. [13].

The corresponding time period to which our estimate refers was computed using the following formula: T = ∑(T(i)*B(i))/∑B(i), where T = the point time location of the global estimate, T(i) = the time location of the estimate for each age group and B(i) = the exposing units of each age group [12]. We carried out stratified analysis based on respondents’ sex, age, and location to see the association with the outcome measure.

Finally, we assessed differences and reduction in MMR in Sidama Regional State and in the districts included in this study using the two maternal mortality estimations methods: the sisterhood method and the 5-year recall of pregnancy and birth outcome household survey [8]. The reference period for the sisterhood method MMR estimation was 2010 whereas the reference period for 5-year recall of pregnancy and child birth outcome MMR estimation was from July 2014-June 2019.

### Ethical approval

We obtained the ethical approval for this study from institutional review board (IRB/015/11) of Hawassa University College of Medicine and Health Sciences and Regional Ethical Committee of Western Norway (2018/2389/REK vest). We got support letter from the Sidama Regional Health Bureau (formerly known Sidama Zone Health Department), and from the district health offices and *kebeles*. Informed (thumb print and singed) consent was obtained from the respondents. We removed the identifiers of the respondents during data entry and analysis.

## Result

Table 1 shows the background characteristics of the respondents. We conducted an interview with 17,444 men and women 15-49 years of age residing in 8880 households. Seventy (0.4%) respondents had incomplete information and were excluded from the analysis. There was no reported death of sisters from the 70 respondents with incomplete information. The final analysis included 17,374 (99.6%) respondents: 8,884 (51.1%) men and 8,490 (48.9%) women. The mean age of the respondents was 29.3 years (SD=6.8), 20.5% had no formal education and 15.2% had attended high school or higher education.

**Table 1.** Characterstics of respondents of maternal mortality survey using the sisterhood method, in Sidama Regional State, southern Ethiopia, 2020

| Characterstics of respondents | Number of respondents | Percentage |
| --- | --- | --- |
| District (Woreda) |  |  |
| Aleta Chuko | 3722 | 21.4 |
| Aleta Wondo | 3929 | 22.6 |
| Aroresa | 2530 | 14.6 |
| Daela | 990 | 5.7 |
| Hawassa Zuriya | 3925 | 22.6 |
| Wondogenet | 2278 | 13.1 |
| Sex |  |  |
| Men | 8490 | 48.9 |
| Women | 8884 | 51.1 |
| Educational level |  |  |
| No formal education | 3557 | 20.5 |
| Grade 1-4 | 4397 | 25.3 |
| Grade 5-8 | 6776 | 39.0 |
| High school or above | 2644 | 15.2 |
| Age group |  |  |
| 15-19 | 706 | 4.1 |
| 20-24 | 3267 | 18.8 |
| 25-29 | 4892 | 28.1 |
| 30-34 | 4216 | 24.3 |
| 35-39 | 2530 | 14.6 |
| 40-44 | 1221 | 7.0 |
| 45-49 | 542 | 3.1 |

Table 2 shows maternal mortality estimates by 5 year age groups. The 17,374 respondents reported a total of 64,387 maternal sisters who had reached 15 years or older, and on average each respondent reported 3.7 sisters. Of the 64,387 reported sisters who had reached 15 years or more, 2,402 (3.7%) sisters had died. From those who had died, 776 (32.3%) were pregnancy related deaths. The total LTR of maternal death was 3.2% i.e. 1 in 31 women 15-49 years old dies due to maternal cause. Using total fertility rate of 5.2, the estimated MMR for the study area was 623 (95% CI: 573-658) per 100,000 LB. The approximate time reference for the MMR estimate was the year 2010, corresponding 10 years prior to data collection.

**Table 2.**
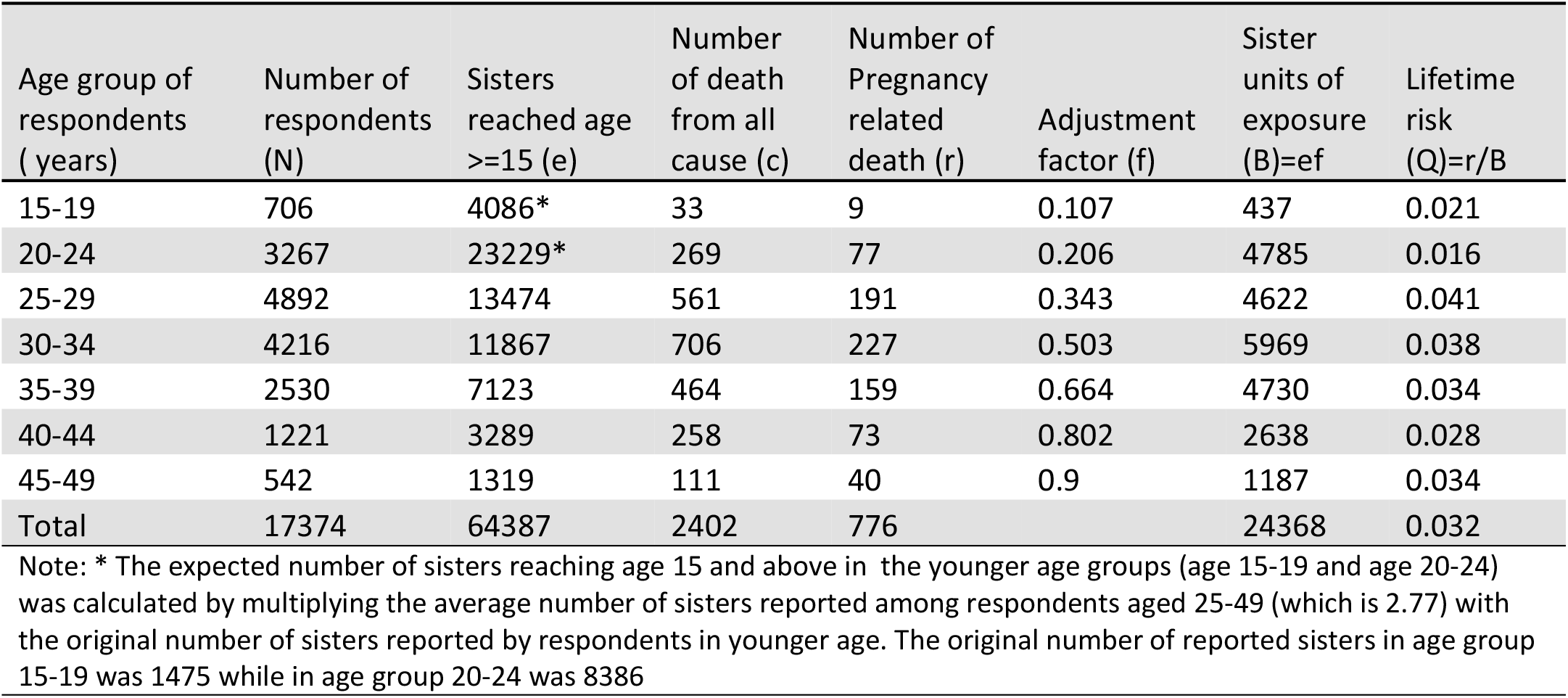
Estimate of lifetime risk of maternal death using the sisterhood method referring to 2010, in Sidama Regional State, southern Ethiopia, 2020

| Age group of respondents ( years) | Number of respondents (N) | Sisters reached age $\geq 15$ (e) | Number of death from all cause (c) | Number of Pregnancy related death (r) | Adjustment factor (f) | Sister units of exposure (B)=ef | Lifetime risk (Q)=r/B |
| --- | --- | --- | --- | --- | --- | --- | --- |
| 15-19 | 706 | 4086* | 33 | 9 | 0.107 | 437 | 0.021 |
| 20-24 | 3267 | 23229* | 269 | 77 | 0.206 | 4785 | 0.016 |
| 25-29 | 4892 | 13474 | 561 | 191 | 0.343 | 4622 | 0.041 |
| 30-34 | 4216 | 11867 | 706 | 227 | 0.503 | 5969 | 0.038 |
| 35-39 | 2530 | 7123 | 464 | 159 | 0.664 | 4730 | 0.034 |
| 40-44 | 1221 | 3289 | 258 | 73 | 0.802 | 2638 | 0.028 |
| 45-49 | 542 | 1319 | 111 | 40 | 0.9 | 1187 | 0.034 |
| Total | 17374 | 64387 | 2402 | 776 |  | 24368 | 0.032 |

Note: \* The expected number of sisters reaching age 15 and above in the younger age groups (age 15-19 and age 20-24) was calculated by multiplying the average number of sisters reported among respondents aged 25-49 (which is 2.77) with the original number of sisters reported by respondents in younger age. The original number of reported sisters in age group 15-19 was 1475 while in age group 20-24 was 8386

To estimate MMR for the most recent period, we repeated the procedure in Table 2 stratifying the participants in three age group: 15-29, 30-39 and 40-49 (Table 3 A). The MMR among participants in the 15-29 age group was 545 (95% CI: 475-601) per 100, 000 LB, reflecting 7 years before data collection. For respondents in the 30-39 age group, the MMR was 703 (624-760) per 100, 000 LB, reflecting a period 11 years before the study. For respondents 40 years or more, the MMR was 564 (455-660) per 100, 000 LB, reflecting a period 15 years before data collection.

**Table 3A.** Maternal mortality estimate and reference period by age group using the sisterhood method, in Sidama Regional State, southern Ethiopia, 2020

| Age group | 15-29 | 30-39 | 40-49 | Total |
| --- | --- | --- | --- | --- |
| Respondents | 8865 | 6746 | 1763 | 17374 |
| Sisters 15y+ | 40789 | 18990 | 4608 | 64387 |
| Sisters 15y+ died | 863 | 1170 | 369 | 2402 |
| Sisters 15y+ died maternal | 277 | 386 | 113 | 776 |
| Sisters exposed to risk | 9844 | 10699 | 3825 | 24368 |
| Lifetime risk | 0.028 | 0.036 | 0.029 | 0.032 |
| MMR* | 545 | 703 | 564 | 623 |
| 95% CI for MMR | 476-601 | 624-760 | 455-660 | 573-658 |
| Estimated years back | 7.4 | 10.6 | 15.3 | 10.2 |
| Note: * Maternal mortality ratio per 100, 000 LB, CI: Confidence Interval |  |  |  |  |

Table 3B shows the life time risk of maternal death and MMR stratified by male and female respondents. There was no statistically significant difference among male and female respondents.

**Table 3B.** Maternal mortality estimate by sex of respondents, using the sisterhood method, in Sidama Regional State, southern Ethiopia, 2020

|  | Male | Female | Total |
| --- | --- | --- | --- |
| Respondents | 8490 | 8884 | 17374 |
| Sisters 15y+ | 26087 | 38485 | 64387 |
| Sisters 15y+ died | 1276 | 1126 | 2402 |
| Sisters 15y+ died maternal | 397 | 379 | 776 |
| Sisters exposed to risk | 12830 | 11574 | 24368 |
| Lifetime risk | 0.031 | 0.033 | 0.032 |
| MMR* | 604 | 643 | 623 |
| 95% CI for MMR | 538-654 | 572-697 | 573-658 |

Note: \* Maternal mortality ratio per 100,000 live birth, CI: Confidence Interval

Table 3C shows stratified analysis of maternal mortality estimates by the districts of respondents. The MMR in Aroresa district was significantly higher than all other study districts in the region; MMR: 1210 (95% CI: 1027-1318) per 100,000 LB.

**Table 3C.**
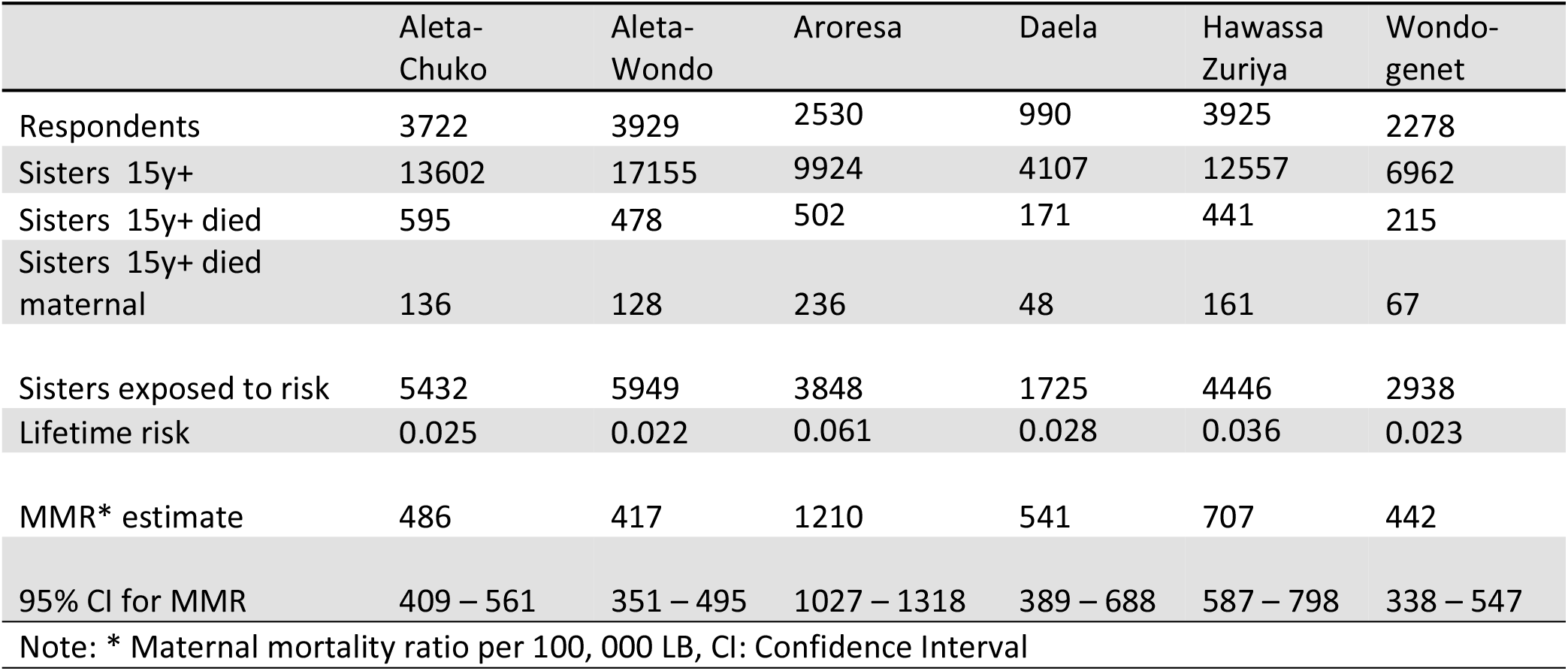
Maternal mortality estimate by district of respondents using the sisterhood method, in Sidama Regional State, southern Ethiopia, 2020

|  | Aleta-Chuko | Aleta-Wondo | Aroresa | Daela | Hawassa Zuriya | Wondogenet |
| --- | --- | --- | --- | --- | --- | --- |
| Respondents | 3722 | 3929 | 2530 | 990 | 3925 | 2278 |
| Sisters 15y+ | 13602 | 17155 | 9924 | 4107 | 12557 | 6962 |
| Sisters 15y+ died | 595 | 478 | 502 | 171 | 441 | 215 |
| Sisters 15y+ died maternal | 136 | 128 | 236 | 48 | 161 | 67 |
| Sisters exposed to risk | 5432 | 5949 | 3848 | 1725 | 4446 | 2938 |
| Lifetime risk | 0.025 | 0.022 | 0.061 | 0.028 | 0.036 | 0.023 |
| MMR* estimate | 486 | 417 | 1210 | 541 | 707 | 442 |
| 95% CI for MMR | 409 – 561 | 351 – 495 | 1027 – 1318 | 389 – 688 | 587 – 798 | 338 – 547 |

Note: \* Maternal mortality ratio per 100, 000 LB, CI: Confidence Interval

Table 4 describes the estimated MMR by two different methods. The sisterhood method refers to around 10 years before the study and showed the overall MMR of 623/100,000 LB in Sidama Regional State. The survey with 5yr recall found 419/100,000 LB with slightly overlapping confidence intervals. Some districts had lower MMR in the survey of 5yr pregnancy recall than the sisterhood estimates reflecting 10 years back, whereas others like Aroresa had similar estimates in both.

**Table 4.** Estimates of maternal mortality ratio by two methods in Sidama Regional State, southern Ethiopia, 2020.

| MMR using 5 year recall of<br>pregnancy and birth outcome* |  |  |  | MMR using the indirect sisterhood method** |  |  |  |  |
| --- | --- | --- | --- | --- | --- | --- | --- | --- |
| District | Live<br>birth | Maternal<br>death | MMR (95% CI) | Number of<br>respondents | Pregnancy<br>related<br>deaths | Sister<br>units<br>exposed<br>to risk | LTR | MMR (95% CI) |
| Aleta Chuko | 2265 | 5 | 263 (58-467) | 3722 | 136 | 5432 | 0.025 | 486 (409-561) |
| Aleta Wondo | 2285 | 12 | 525 (207-842) | 3929 | 128 | 5949 | 0.022 | 417 (351-495) |
| Aroresa | 1694 | 21 | 1142 (693-1591) | 2530 | 236 | 3848 | 0.061 | 1210 (1027-1318) |
| Daela | 716 | 4 | 641 (77-1358) | 990 | 48 | 1725 | 0.028 | 541 (389-688) |
| Hawassa- Zuriya | 2237 | 2 | 114 (24-251) | 3925 | 161 | 4446 | 0.036 | 707 (587-798) |
| Wondogenet | 1405 | 4 | 258 (8-508) | 2278 | 67 | 2938 | 0.023 | 442 (338-547) |
| Sidama Region | 10602 | 48 | 419 (260-577) | 17374 | 776 | 24338 | 0.032 | 623 (573-658) |

Note: \*Reference year: July 2014-June 2019, \*\* Reference year: 2010, Abbreviations: MMR: maternal mortality ratio per 100,000 live birth, LTR: life time risk for maternal death, CI: confidence interval

## Discussion

### Principal findings

By incorporating the sisterhood method in a household survey of pregnancy and birth outcomes, we found a lifetime risk of maternal death of 3.2 %, with a corresponding MMR of 623 per 100,000 LB; the time reference was 2010. The remote district (Aroresa) had significantly higher MMR. Sub analysis of MMR based on the respondents’ age showed that participants 15-29 years of age had a MMR; 545 per 100, 000 LB, reflecting 7 years before data collection.

MMR estimated by a household survey that used a 5-year recall of pregnancy found an estimated MMR of 419 per 100,000 LB and the sisterhood method referring to 10 years before the study found 623 per 100,000 LB, slightly overlapping confidence intervals. Districts located distant from the centre with poor infrastructure and inadequate skilled health personnel did had similar MMR by both methods and seem to have persistently high MMR. Whereas, districts located near to the centre with good infrastructure and adequate skilled health personnel have lower estimated MMR by the 5-year recall than by the sisterhood method.

### Strengths and weaknesses of the study

To the best of our knowledge, this is the first study describing maternal mortality estimates employing the sisterhood method in Sidama Regional State, southern Ethiopia. The study was conducted using large and representative sample of the region that demonstrated district level variations. By assessing the MMR of the sisterhood method and the estimates from household maternal mortality survey that used a 5-year pregnancy and birth outcomes [8], this study identified variations in MMR reduction in different geographical areas of the region.

This study had some limitations similar to other studies employed the sisterhood method. The maternal mortality estimates we reported refer a period around 10 years before data collection; thus we were not able to show recent estimates using this method.

There might be over-reporting of deaths. Over reporting of deaths may arise due to inclusion of deaths that occurred beyond six weeks of end of pregnancy or cases which were not related to pregnancy. Multiple counting could also be the reason for over-estimation of maternal deaths using the sisterhood method.

To minimize over-reporting, we used data collectors who were familiar with language and culture of the study population. As a result of familiarity with local context, the data collectors supported participants in identifying pregnancy state of deceased sisters and time of their deaths. In addition, in households where more than one eligible participants born to the same mother were found, we chose one of them by a lottery method which minimized multiple counting. Despite our effort, there might be over-reporting in our study.

Underreporting could also be another limitation for this study. Deaths occurring at early stage of pregnancy due to abortion or ectopic pregnancy and deaths of women not in marital union might not be reported.

We did not have information on place of living for the sisters in the cohort. We used the respondents’ residences as proxy for the sisters’ location. Some sisters might have moved to other districts. Our study also lack information on age at death of respondents’ sisters but used respondents’ age to observe recent deaths and show patterns of deaths across age groups. Age of the respondents may not closely reflect the age of their sisters.

### Maternal mortality ratios and variations using the sisterhood method

We found high MMR in the study area with time reference in 2010. Our result is significantly higher than the findings of national DHS [6], which reported a MMR 412 per 100,000 LB (time reference, 2009-2016). The DHS study uses the direct sisterhood method which included detailed information on age at death, year of deaths and years since the death occurred which contributed for precise maternal mortality estimates [15]. However, in our study, we used the indirect sisterhood method in which such detailed information is not included. The inherent weakness of the indirect sisterhood method lacking exact information on age at death, year of death and years since the death occurred may overestimate the deaths[14].

The estimate from our study was also higher than a study from Kersa Health and Demographic Surveillance Site (HDSS) [16], which estimated a MMR of 396 per 100,000 LB for the year 2010. The variation could be explained by the differences of methods the two studies used for MMR estimation, access to maternal health care and differences in documentation of maternal deaths. The Kersa’s study employed direct method of maternal mortality estimation, whereas we used the indirect sisterhood method which might have caused over reporting of maternal deaths [11]. Mothers in Kersa DHSS might have better access to health related information and improved maternal health service utilization due to DHSS activities which in turn might have contributed for reduction of maternal deaths in the area. In addition, in Kersa’s DHSS there also might be improved vital registration system that supported accurate documentation of maternal deaths.

A study from Gamo-Gofa south-west Ethiopia, using the sisterhood method with a time reference of 1998 found a MMR 1667 per 100,000 LB, which is higher than our study [11] but carried out a decade before our study. The government has implemented many interventions since that study to improve universal health coverage on maternal health services which might have contributed for a reduction of maternal mortality in our area [1].

In our study, we found similar maternal mortality estimates both from men and women respondents. This shows that the information obtained from men siblings is as valid as women siblings in estimating maternal mortality using the indirect sisterhood method. This is in agreement with report from south-west Ethiopia [11]. A study carried out using DHS data from African countries also found similar findings [17].

This study found high maternal mortality estimates among the respondents in the 30-39 age group; MMR: 703 per 100,000 LB compared to participants in 15-29 years of age. Our finding is different from findings reported from Nigeria where they found the highest MMR among the respondents in the 15-29 age group [10]. Though we did not have information on age of the diseased sisters (only respondents), the difference in age at first marriage might have contributed for the variations observed in two settings. In Ethiopia, the average age at first marriage was reported to be 17 [6] while 13 years was reported from Mali [9].

Our study has also shown significant variations in MMR by districts of the respondents. This is in agreement with the findings of study conducted in northern Nigeria and Mali [9, 10] where they reported that maternal mortality differed by local government areas and villages respectively. The areas with high maternal mortality in those studies were characterized by remoteness and problematic access to health facilities. Previously, studies have used the place of the respondents as a proxy location for the sisters of the respondents to describe the differences in deaths based on the location of the respondents [9, 10].

The high MMR observed in Aroresa districts (the remote district) could be explained by poor road facilities and difficult topography that might hamper access to health services. Aroresa district is situated 181 km away from the regional centre [18]. The distance from the centre may affect availability of services during emergency situation. Weak emergency obstetric care compounded with lack of adequate and skilled personnel might have also contributed for the high maternal mortality in the district. Low utilization rate of maternal health services could also explain the high MMR in Aroresa district, and a paper from this area in 2018 reported institutional delivery in 38% and use of contraceptives in 49% [18].

### Maternal mortality reduction in Sidama Regional State

A study from India reported there existed regional and district level variations in maternal mortality reduction across the country [19]. In our study of Hawassa Zuriya district (the central district), the sisterhood study reflecting 10 years back gave a MMR of 707 and 5y recall gave 114 per 100.000 LB. The apparent reduction in MMR in this district could be attributed to improved access to obstetric care during emergency situation as the district is located near to the referral centre at the regional capital Hawassa [8] with good infrastructure and road facility. Moreover, the district is characterized by having relatively high doctor to population ratio and midwife to population ratio which might have improved access to skilled obstetric care [8]. An Indonesian study demonstrated that investment in doctors and hospitals would significantly reduce the MMR [20]. Contrary to Hawassa Zuriya district (the central district), in Aroresa district (the remote district), the two studies found high MMR with no reduction of MMR over the past years.

### Validity of maternal mortality measurements

We incorporated the sisterhood questions to the ongoing maternal mortality household survey that used a 5-year recall of pregnancy and birth outcomes to determine the MMR. We added 4 questions of the indirect sisterhood to the main study questionnaire and collected the data from respondents residing in the same households sampled for the main study using the same enumerators. There was no major cost and logistics incurred due to inclusion of the sisterhood questions to the main household survey questionnaire. However, the main household study incurred costs in terms of time, logistics and money. Despite the costs incurred, such studies provide important information on magnitude and variations of MMR which is useful for program planning, priority setting and resource allocation at lower administrative level.

We considered the size of the sample and sampling techniques while conducting the studies using the two maternal mortality estimation methods in order to precisely estimate the MMR in the region. Hence, large number of participants were included in both 5-year recall of pregnancy and birth outcome household survey and the sisterhood study. In 5-year recall of pregnancy and birth outcome household survey, we registered 10602 LB and 48 maternal deaths in 8880 households visited. Our aim was to find 66 maternal deaths with MMR; 412

(95% CI: 324-524) per 100,000 LB. The MMR after the study was 419 (95% CI: 260-577) which is within the 95% CI we anticipated initially [8]. For the sisterhood study, we had an interview with 17444 siblings. This sample size was above the recommended 13,000 siblings for the indirect sisterhood study in settings with similar magnitude of maternal mortality [13].The samples for both studies were selected using probability sampling and multistage cluster sampling technique was employed to select the study participants.

The MMR we found using the sisterhood method was higher than the findings from other studies in the same time reference [6]. Over-estimation of MMR from studies that used the sisterhood method have been documented [11]. The MMR obtained from the household survey was comparable with other national figures [6, 7].

Recall bias is a potential limitation both in the household survey study and the sisterhood study as the conclusions of the studies were based on the information obtained from families memorizing past events of maternal deaths. We believe that the recall bias is low in 5-year recall of pregnancy and birth outcome study than the sisterhood as the deaths were occurred in the recent years and the interviews were conducted with close relatives who lived together with the deceased mother.

Under-estimation could also be another limitation for both studies. Stigma of pregnancies of unmarried girls may lead to under-reporting. Maternal deaths occurring at early stage of pregnancy due to abortion or ectopic pregnancy may be missed and regarded as non-maternal deaths within the community.

### Policy and clinical implications

Persistently high maternal mortality in districts found in remote area, far away from central city with poor infrastructure and inadequate skilled health workers calls for the attention from the government and the importance of instituting interventions tailored to the local context to improve poor infrastructure and shortage of skilled health personnel in Sidama Region State.

Analysis of results from recent maternal mortality estimation using the 5-year recall of pregnancy and birth outcome household survey with the sisterhood method suggests the possibility of monitoring maternal mortality reduction at regional and district level using the two estimation methods.

## Conclusion

This study used two methods for estimating maternal deaths in Sidama, Ethiopia, one referring to last 5 years and the other 10 years back. The central districts have a lower MMR than before, but peripheral districts still have high MMR. The Sidama Regional Health Bureau and respective district health offices should implement interventions tailored to the local context to address variations and accelerate reductions in MMR.

## Data Availability

All relevant data are within the manuscript

## Acknowledgements

We are grateful to the participants of the study for their time and information. Our special gratitude goes to Sidama Regional Health Bureau, respective district health offices and *kebele* administrations in Ethiopia for their support and permission to conduct the study.

## Authors’ contributions

AZK, BL and SGH conceived and designed the experiments. AZK, BL and SGH performed the experiments. AZK analysed the data. BL, AGT and SGH supervised and provided mentorship. AZK drafted the article. All authors contributed to the writing, reviewed the article and approved the final version of the manuscript.

## References

1. Ayele AA, Tefera YG, East L. Ethiopia’s commitment towards achieving sustainable development goal on reduction of maternal mortality: There is a long way to go. Women’s Health. 2021; 17. doi: 10.1177/17455065211067073 PMID: 34913391

2. World Bank, World Health Organization. Global Civil Registration and Vital Statistics: Scaling up Investment Plan 2015-2024. World Bank Group. 2014.

3. Abay ST, Gebre-Egziabher AG. Status and associated factors of birth registration in selected districts of Tigray region, Ethiopia. BMC Int Health Hum Rights. 2020; 20 (20). doi: 10.1186/s12914-020-00235-x PMID:32727474

4. Tesfaye G, Loxton D, Chojenta C, Assefa N, Smith R. Magnitude, trends and causes of maternal mortality among reproductive aged women in Kersa health and demographic surveillance system, eastern Ethiopia. BMC Women’s Health. 2018; 18(198). doi: 10.1186/s12905-018-0690-1 PMID: 30518368

5. Legesse T, Abdulahi M, Dirar A. Trends and causes of maternal mortality in Jimma University Specialized Hospital, southwest Ethiopia: a matched case-control study. Int J Women’s Health. 2017;9: 307–313. doi: 10.2147/IJWH.S123455 PMID: 28496370

6. Central Statistical Agency Ethiopia:The DHS Program ICF Rockville Maryland, USA. Demographic and Health Survey 2016 Final Report.2017.

7. WHO, UNICEF, UNFPA, WORLD BANK, UNPD. Trends in Maternal Mortality:2000-2017. WHO, UNICEF, UNFPA, UNDP and The World Bank estimates. 2019.

8. Aschenaki Zerihun Kea, Bernt Lindtjorn, Achamyelesh Gebretsadik, Hinderaker SG. Variation in maternal mortality in Sidama Regional State, southern Ethiopia: A population based cross sectional household survey. medRxiv 277635 [preprint] [posted 2022 July 14]. Available from: https://medrxiv.org/cgi/content/short/2022.07.14.22277635v1

9. Aa I, Grove MA, Haugsja AH, Hinderaker SG. High maternal mortality estimated by the sisterhood method in a rural area of Mali. BMC Pregnancy Childbirth. 2011; 11(56). doi: 10.1186/1471-2393-11-56 PMID: 21812951

10. Sharma V, Brown W, Kainuwa MA, Leight J, Nyqvist MB. High maternal mortality in Jigawa State, Northern Nigeria estimated using the sisterhood method. BMC Pregnancy Childbirth. 2017; 17(163). doi: 10.1186/s12884-017-1341-5 PMID: 28577546

11. Yaya Y, Lindtjorn B. High maternal mortality in rural south-west Ethiopia: estimate by using the sisterhood method. BMC Pregnancy Childbirth. 2012; 12(136). doi: 10.1186/1471-2393-12-136 PMID: 23176124

12. Graham W, Brass W, Snow RW. Estimating maternal mortality: the sisterhood method. Stud Fam Plann. 1989; 20(3):125-35. PMID: 2734809

13. Hanley JA, Hagen CA, Shiferaw T. Confidence intervals and sample-size calculations for the sisterhood method of estimating maternal mortality. Stud Fam Plann. 1996; 27(4):220-227. PMID: 8875734

14. World Health Organization, United Nations Children’s Fund. The Sisterhood Method for Estimating Maternal Mortality: Guidance notes for potential users. 1997.

15. N. Rutenberg, J.M. Sullivan. Direct and indirect estimates of maternal mortality from the sisterhood method. IRD/Macro International Incl. 1991.

16. Tesfaye G, Loxton D, Chojenta C, Assefa N, Smith R. Magnitude, trends and causes of maternal mortality among reproductive aged women in Kersa health and demographic surveillance system, eastern Ethiopia. BMC Women’s Health. 2018; 18(198). doi: 10.1186/s12905-018-0690-1 PMID: 30518368

17. Merdad L, Hill K, Graham W. Improving the measurement of maternal mortality: the sisterhood method revisited. PLoS One. 2013; 8(4):e59834. doi: 10.1371/journal.pone.0059834 PMID: 23565171

18. Limaso AA, Dangisso MH, Hibstu DT. Neonatal survival and determinants of mortality in Aroresa district, Southern Ethiopia: a prospective cohort study. BMC Pediatr. 2020; 20(33). doi: 10.1186/s12887-019-1907-7 PMID: 31987037

19. Srinivas Goli, Parul PuriI, Pradeep S. Salve, Saseendran Pallikadavath, K.S. James. Estimates and correlates of district-level maternal mortality ratio in India. PLOS GLOBAL PUBLIC HEALTH 2022;2(7). doi: 10.1371/journal.pgph.0000441

20. Cameron L, Contreras Suarez D, Cornwell K. Understanding the determinants of maternal mortality: An observational study using the Indonesian Population Census. PLoS One. 2019; 14(6):e0217386. doi: 10.1371/journal.pone.0217386 PMID: 31158243

